# Genetic Diagnosis of Facioscapulohumeral Muscular Dystrophy Type 1 Using Rare Variant Linkage Analysis and Long Read Genome Sequencing

**DOI:** 10.1101/2023.06.05.23290975

**Authors:** Kun Li, Daniel Quiat, Fei She, Yuanwei Liu, Rong He, Alireza Haghighi, Fang Liu, Rui Zhang, Steven Robert DePalma, Ying Yang, Wen Wang, Christine E. Seidman, Ping Zhang, Jonathan G. Seidman

**Author notes:** These authors contributed equally to this work (Kun Li, Daniel Quiat).

## Abstract

Facioscapulohumeral dystrophy type 1 (FSHD1) is a progressive, debilitating skeletal myopathy that requires a multimodal approach for complete molecular characterization of pathogenic genotypes. Here, we report genomic analyses of a family with suspected facioscapulohumeral dystrophy type 1 (FSHD1). We first performed short read genome sequencing, followed by parametric linkage analysis using rare variants to map the disease locus to a single 1.7 MB interval on chromosome 4q35.2 with a LOD score of 3.2. We then used ultra-long read genome sequencing as a single molecular test to genotype a pathogenic FSHD allele containing a 4qA permissive haplotype and 5 KpnI repeat units at the D4Z4 locus. These results demonstrate that genome-wide rare variant-based linkage analysis is a powerful tool for mapping disease loci in families and ultra-long read genome sequencing is capable of genotyping pathogenic FSHD1 alleles.

## Introduction

Facioscapulohumeral dystrophy type 1 (FSHD1, OMIM:158900) is an autosomal dominant myodystrophy with progressive and regional skeletal muscle involvement and variable disease penetrance^1,2^. The pathogenic genetic mechanism of FSHD1 is epigenetic de-repression of the *DUX4* gene encoded within macrosatellite tandem repeats at the D4Z4 locus in the subtelomeric region of chromosome 4q.^3^ Unaffected individuals carry 11-100 highly similar 3.3-kb KpnI repeat units at the D4Z4 locus, each containing one copy of entire open reading frame (ORF) of the gene encoding the transcriptional regulator *DUX4*, which is normally expressed during embryonic development and restricted to testis and thymus in somatic tissues. Affected individuals carry an allele with 1-10 KpnI repeat elements and a permissive 4qA haplotype containing a polyadenylation signal within the adjacent the pLAM sequence, that together result in aberrant expression of *DUX4*, which is toxic to skeletal muscle cells^1,4-7^.

Molecular diagnosis of FSHD1 is routinely made by Southern blotting, which requires a substantial amount of input DNA and often yields inconclusive results in up to 23% of cases^8^. More recently, assessment of D4Z4 repeat length by whole genome optical mapping and molecular combing have been used for clinical genetic testing^9,10^. Sequencing of the pathogenic allele using short-read genome sequencing or Sanger sequencing is technically difficult due to low complexity repeats with high GC content and the existence of a highly homologous subtelomeric region on chromosome 10q26 ^11^. Recently developed long-read sequencing methods, such as Oxford Nanopore Technologies (ONT) sequencing platform, are capable of sequencing single DNA molecules that are kilobases in length, enabling superior sequencing and assembly of repetitive regions of the human genome ^12^. This, and other long read genome sequencing platforms are promising alternatives for a single molecular test for FSHD. These long-read genome sequencing methods can detect pathogenic alleles by sequencing the complete D4Z4 repeat array in a single read enabling quantification of KpnI repeats and detection of the permissive 4qA disease haplotype. In addition, pathogenic variants in the gene *SMCHD1* that result in the closely related Facioscapulohumeral dystrophy type 2 (FSHD2, OMIM: 158901) phenotypes can be evaluated.

Here, we use a combination of short and long read genome sequencing in a multiplex kindred to map and diagnose FSHD1 respectively.

## Results

### Clinical phenotyping of the affected kindred

We identified a five generation kindred with hereditary myopathy (Figure 1A). The proband was a male in his 30s who developed progressive muscle weakness between 10-20 years of age, and went on to develop muscular atrophy in the shoulder, back, and proximal extremities and scapular winging. The proximal muscle strength of his bilateral upper limbs was measured as grade 4,^13^ his distal muscle strength was normal, and neurological examination was overall negative. There was no obvious reduced facial expression, facial movements, and facial diplegia. He reported being able to continue performing physical activities of daily living without limitations. Detailed family history revealed multiple family members with a similar phenotype of symmetrical or asymmetrical, progressive, proximal muscle atrophy and weakness that demonstrated autosomal dominant mode of inheritance (Figure 1A, additional phenotypic details redacted but available from the authors upon request). The median age at onset was 26 (IQR: 22.5, 40) years and thirty eight percent of patients were male. Thirteen percent of affected individuals had obvious lower limb weakness. A clinical diagnosis of familial myopathy was made without definitive diagnosis.

**Figure 1.**
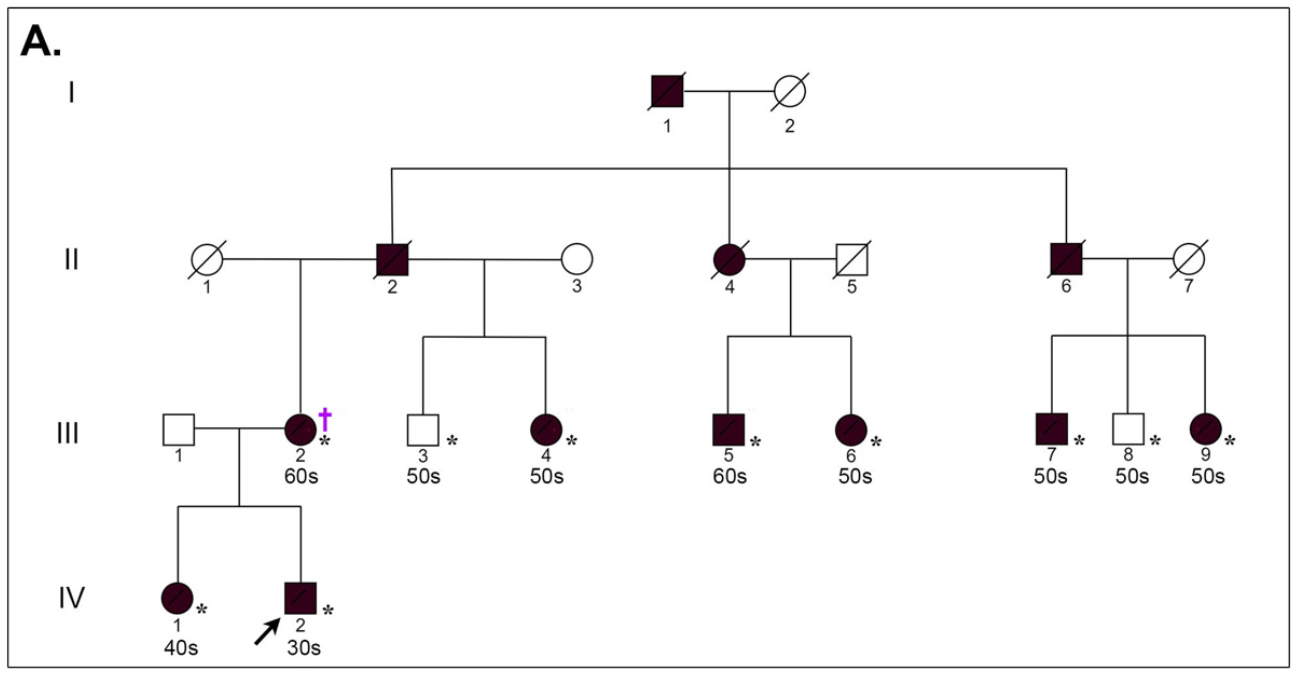
Pedigree of a kindred with suspected FSHD. (**A**) A multiplex kindred with suspected FSHD. The proband (IV-2) is indicated by a black arrow. Asterisks(*) denote individuals who underwent Illumina short read genome sequencing and dagger (†) denotes evaluation by ultra-long nanopore sequencing. Males are represented by squares, females by circles, and affected members by black symbols. Deceased individuals are marked by a diagonal line. Age by decade is noted below individuals who underwent short read genome sequencing.

### Rare-variant linkage mapping of the disease locus

Exome sequencing for the proband (IV-2) and his mother (III-2), including analysis of established and candidate myopathy genes, did not identify any candidate causal variants. We next performed genome sequencing on eight affected and two unaffected individuals (Figure 1A, asterisk) and did not identify any rare damaging SNPs, indels, genomic structural variants, or splice site variants affecting established or candidate myopathy genes shared across affected individuals. We then sought to leverage genome sequencing data to map the candidate disease locus, specifically the ability to genotype rare SNPs that could mark the haplotype containing the disease allele. Based on the autosomal dominant mode of inheritance, we hypothesized that the haplotype containing the disease allele could be mapped by identifying ultra-rare heterozygous variants shared by all affected individuals and absent from unaffected relatives. We identified 15417 genome-wide ultra-rare heterozygous biallelic SNPs with an allele frequency of less than 1×10^−4^ in East Asian subjects in the genome aggregation database (gnomAD). We performed parametric linkage analysis with these 15417 ultra-rare SNP markers and parameters of an autosomal dominant model and estimated penetrance of 90%. Only a single haplotype on the terminus of chromosome 4q35.2, marked by 4 rare biallelic SNPs (Supplemental Table 1), was shared by all affected family members and was absent from unaffected controls, and had a LOD score of 3.2 suggesting evidence of genetic linkage (Figure 2A). Evaluating the haplotypes more broadly, we noted two cross-over events that facilitated mapping of the locus (Figure 2B).

**Figure 2.**
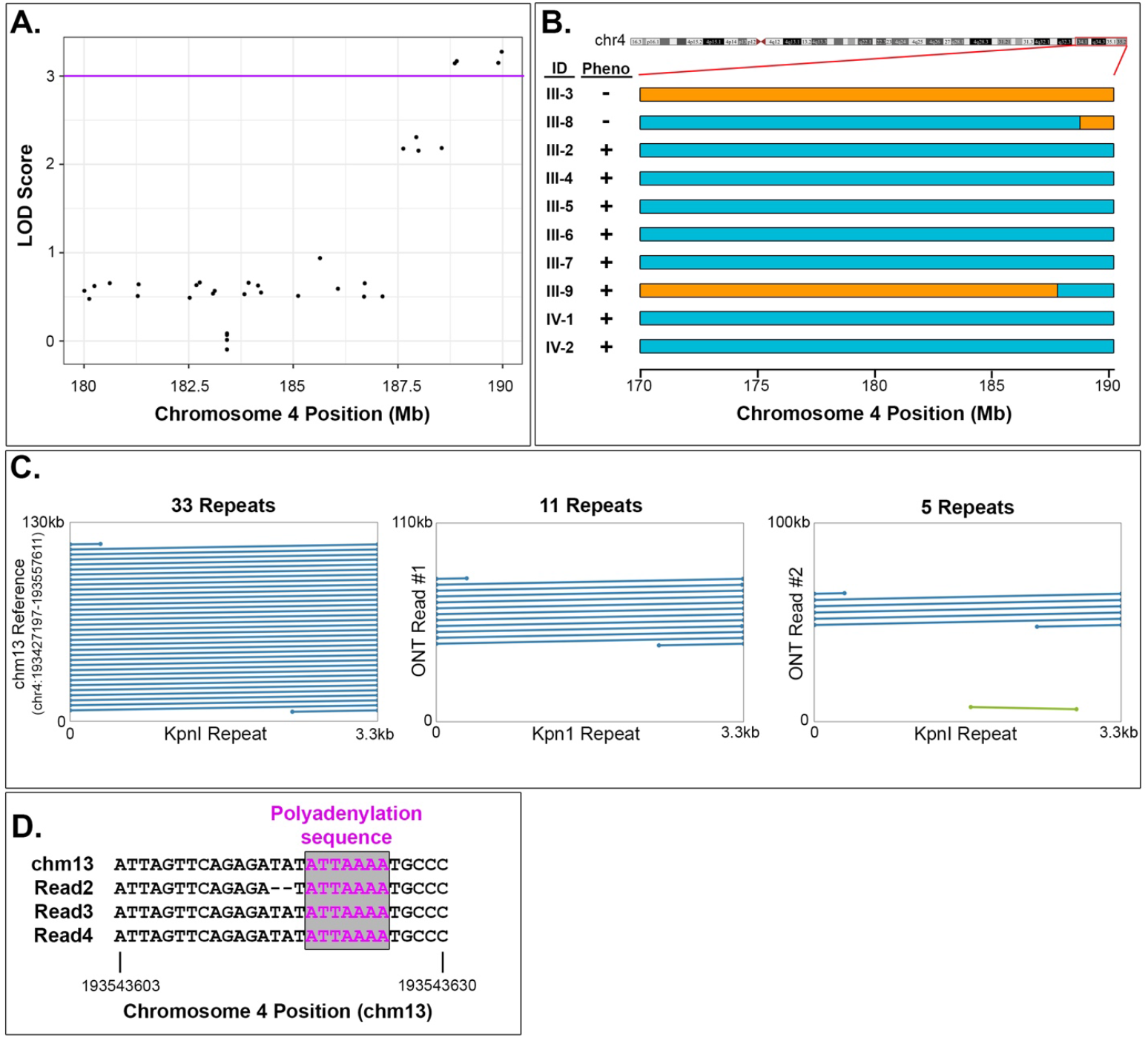
Genetic mapping and ultra-long read sequencing of the pathogenic allele. (**A**) Jitter plot of the logarithm of the odds (LOD) score for genetic linkage between rare biallelic SNPs with a population allele frequency of less than 1×10^−4^ in East Asian individuals in the gnomAD database indicates linkage at the terminus of chromosome 4q35.2. (**B**) Schematic of the haplotypes marked by shared rare variants in affected (Pheno ‘+’) and unaffected (Pheno ‘-’) individuals demonstrates the crossing over events that demarcate the pathogenic haplotype. Disease associated haplotype is depicted in cyan and other haplotypes are in orange. (**C**) Sequence alignment of the 3.3kb KpnI repeat sequence with the chm13 reference genome demonstrates the presence of 33 repeats in D4Z4 region of the reference sequence. Alignment of ultra-long Oxford Nanopore reads demonstrates the presence of a non-pathogenic allele (‘Read 1’) with 11 KpnI repeats and a pathogenic allele (‘Read 2’) with 5 KpnI repeats in an affected individual. (**D**) Multiple sequence alignment of the chm13 reference genome and sequencing reads containing 5 KpnI repeats (Reads 2-4) demonstrates the presence of the polyadenylation sequence of the 4qA permissive haplotype.

### Molecular Diagnosis of FSHD1 by ultra-long read genome sequencing

The region on chromosome 4q35.2 identified by linkage analysis has previously been associated with FSHD1 and matched the phenotype of the affected individuals in the kindred.^14^ Clinical genotyping for FSHD1 pathogenic alleles requires analysis of 4q35.2 D4Z4 repeat array length and haplotyping by southern blot or other techniques^15^. Given the complex, multimodal protocol for genotyping pathogenic FSHD1 alleles, and previous reports of Oxford Nanopore sequencing to assess D4Z4 repeat array length in BAC clones,^11^ we hypothesized that third-generation ultra-long read genome sequencing could be used to define the FSHD1 allele in this kindred and confirm the diagnosis of FSHD1. We performed ultra-long read Oxford Nanopore genome sequencing in affected individual III-2 (Figure 1A) with a mean read length of 34,910 bp and read length N50 of 62,623 bp (Supplemental Figure 1A). We then aligned reads to the chm13 (v2) reference genome, which unlike the hg38 reference sequence contains a gapless assembly of complex repetitive sequence on 4q35.2. The chm13 reference sequence contains 33 KpnI repeat elements in the D4Z4 array on chromosome 4q35.2 (Figure 2C). We identified 4 sequencing reads that aligned to 4q35.2 and spanned the D4Z4 repeat array. One 110 kilobase (kb) long read contained 11 KpnI repeats (‘Read 1’, Figure 2B), inconsistent with an FSHD1 pathogenic allele that has less than 10 KpnI repeats. Three reads (66kb, 100kb, 160kb in length) contained 5 KpnI repeat elements (‘Reads 2-4’, Figure 2C and Supplemental Figure 2) and the 4qA permissive haplotype with ATTAAA polyadenylation sequence (Figure 2D) consistent with a pathogenic FSHD1 haplotype in an affected individual.

## Discussion

We report characterization of a large kindred with myopathy of unclear etiology exhibiting autosomal dominant inheritance. By employing both short and long read genome sequencing we demonstrate that the disease maps to the FSHD1 locus and that the proband carries a pathogenic allele. Pathogenic FSHD1 alleles contain a contraction of D4Z4 macrosatellite repeat array, requiring genotyping by Southern blot analysis^15^. Some FSHD1 cases are caused by the presence of hybrid alleles, mosaicism and D4Z4 array rearrangements, making the results of standard molecular testing difficult to interpret. Several alternative methods for simplified FSHD genotyping have been proposed. Molecular combing can overcome some of the limitations of southern blotting by providing direct visualization of the D4Z4 repeats on chromosome 4q and 10q, and even the precise size of D4Z4 repeats^10^. Optical genome mapping can also determine the size and haplotype of D4Z4 alleles for FSHD analysis, but the processing steps are complex, particularly with the initial DNA preparation and labeling akin to the complexities of preparing the genomic DNA and performing Southern blot analysis for FSHD^9^. We demonstrate that third generation long-read sequencing can be used to genotype FSHD1 pathogenic alleles. Advantages of sequencing-based genotyping include potential for complete FSHD1 and FSHD2 genotyping in a single assay that can assess genomic structural changes (D4Z4 KpnI repeat number), 4qA/B haplotype status, and damaging variants in *SMCHD1* that are associated with FSHD2. Application of short read sequencing based testing for molecular diagnosis has been limited in certain regions of the human genome, including the 4q35.2 region, due to high homology, repetitive sequence, and GC-rich regions^16^. Furthermore, the human reference sequence builds (hg38 and earlier) do not accurately incorporate these repetitive regions, limiting the application of reference-based sequencing methods.

In our study, we utilized ultra-long read ONT sequencing to obtain a molecular diagnosis in a large kindred with suspected FSHD. We obtained 3 sequencing reads from a pathogenic allele and one read from the non-pathogenic allele spanning the D4Z4 repeat array, with 5 and 11 KpnI repeats respectively, among 756,190 total reads that passed QC. Despite achieving ultra-long read lengths with N50 of 62,623 bp, we only identified 3 reads with the pathogenic allele and one from the non-pathogenic allele. Increasing the overall sequencing depth, use of adaptive sampling to selectively sequence DNA molecules from areas of interest (4q35.2, *SMCHD1* locus, etc), or performing target capture prior to sequencing will likely improve the yield of ultra-long reads available for robust molecular analysis.

In addition to use of long read sequencing to genotype the pathogenic allele, we used rare variants with an allele frequency of less than 1×10^−4^ for linkage analysis and four SNPs were identified with robust evidence of linkage. These markers could be utilized for pre-clinical test for screening children with phenotype unknown, albeit recognizing limitations due to potential recombination events.

In summary, our genome sequencing study of a large kindred with FSHD1 demonstrates the use of genome-wide rare variants-based linkage analyses as a powerful tool for the detection of pathogenic regions in multiplex kindreds, and the potential for ultra-long nanopore sequencing as a genetic testing modality for diagnosis of FSHD1.

## Methods

### Subjects and Ethics Declaration

Our primary cohort consists of eight patients and two unaffected individuals from a large kindred spanning five generations with suspected FSHD recruited at a single center by a single provider. Genomic DNA was extracted from peripheral blood samples of the eight affected and two unaffected family members (Figure 1a). Elements of this study involving human subjects were approved by the Medical Ethics Committee of Beijing Tsinghua Changgung Hospital, Tsinghua University. All participants provided informed written consent before initiating research and data collection, including the use and publication of photographs in an open-access publication. All research was performed in accordance with the principals outlined in the Declaration of Helsinki.

### Clinical assessment

We collected data on ten family members including the proband (VI-2) with their ages, gender, physical examination, investigations, and clinical manifestation details. History-taking, physical examination, and electrocardiogram (ECG) were performed for all included family members. Radiology, electromyography (EMG) and echocardiography were only performed for the proband. Muscle strength is scored according to the Medical Research Council Manual Muscle Testing scale^13^. Facial muscles were assessed by completing specific motions, which include whistling, smiling, puckering lips, puffing out cheeks and burying eyelashes. Limb muscles were evaluated by manual muscle testing. Scapular muscle weakness was confirmed according to the standard as follows: 1) scapular wing when abducting arms; 2) unable to lift arms to head level; 3) obvious scapular muscle atrophy ^17^.

### Histological analyses

Biceps muscle biopsy of the proband were investigated with histological and enzyme histochemical analysis. For histochemical staining, transverse 8 μm cryostat muscle sections were stained with haematoxylin & eosin (H&E), modified gomori trichome (mGT), Nicotinamide adenosine dinucleotide-tetrazolium reductase (NADH-TR), adenosine triphosphatase (ATP), periodic acid Shiff (PAS), cytochrome oxidase (CCO), oil red O (ORO), acid phosphatase (ACP) and non-specific esterase (NES). Immunohistochemistry for CD3, CD4, CD8, CD20, CD68 and C5b-9 was performed in muscle samples.

### Whole exome sequencing (WES)

Blood samples were collected for the proband and his mother. Libraries were prepared according to the protocols of Agilent SureSelect QXT Library Prep Kit (5500-0127), and exome capture was conducted based on the protocols of Agilent SureSelect QXT Target Enrichment for Illumina Multiplexed Sequencing version E0. Library DNA quantity and quality were evaluated by Agilent 2200 TapeStation before hybridization. The enriched libraries on the beads were amplified by polymerase chain reaction (PCR) and cleaned with Ampure XP beads (Agencourt, Boston, MA, USA) according to the SureSelect QXT protocol. Paired-end 2×150 bp sequencing was performed on Illumina NovaSeq 6000 System. Genome Analysis Toolkit (GATK) was used to identify variants. Synonymous variants were filtered and excluded. Deleterious single-nucleotide variants (SNVs) were predicted by PolyPhen-2, SIFT and Clinvar. Variants were assessed according to the ACMG (American College of Medical Genetics and Genomics) guideline ^18^.

### Short Read Genome Sequencing

The NadPrep® EZ DNA Library Preparation Module (for Illumina®) kit was used for library construction, followed by high-throughput, high-depth sequencing on the Illumina Novaseq platform. The genomic DNA is randomly broken into fragments with a length of about 250 bp by endonuclease, and after end repair and A-tail addition, DNA libraries with specific tags are connected to the two ends of the fragments respectively. Finally, PCR amplification of the library was performed. After the library was constructed, the Agilent 2200 Tape Station was used to detect the insert size of the library. qPCR was subsequently adopted to accurately quantify the effective concentration of the library to ensure the quality of the library. Illumina Novaseq platform PE150 sequencing was performed according to the effective concentration of the qualified library and data output requirements.

### Short read sequencing analysis

Reads underwent adapter trimming, then were aligned to the hg38 reference genome using the Burrows-Wheeler Aligner (BWA-MEM) and processed in accordance with Genome Analysis Toolkit (GATK, Broad Institute) Workflow best practices^19^. Briefly, sequence alignments underwent BaseQuality Score Recalibration (BQSR, GATK) and marking of duplicates (MarkDuplicates, Picard), followed by individual level variant calling with HaplotypeCaller (GATK). Individual samples were then joint-genotyped, and variants underwent Variant Quality Score Recalibration (VQSR, GATK). Final SNP and indel calls were filtered with the ‘PASS’ filter from VQSR LOD scoring, read depth (DP) >=10, and genotype quality (GQ) >=20, and subsequently phased with Eagle (v2.4.1) using the 1000 Genomes Project Phase 3 reference panel. To evaluate for genomic structural variants (duplications, deletions, inversions, translocations > 50bp) we ran GATK-SV to with a single sample mode on Terra.

### Rare variant parametric linkage analysis

Under the assumption of an autosomal dominant model based on the pedigree, we used bcftools (v1.10) to filter variant calls for rare heterozygous biallelic SNPs with a frequency in East Asian subjects (EAS) in gnomAD (v3) less than 1E-4. We then used these haplotype-defining SNP markers to perform a parametric linkage analysis using Merlin (v1.1.2) with the following parameters: dominant model, an estimated population allele frequency of 1E-4, and penetrance of 90%.

### Nanopore Sample Preparation

High molecular weight genomic DNA was prepared by the CTAB method and followed by purification with QIAGEN® Genomic kit (Cat#13343, QIAGEN) for regular sequencing, according to the standard operating procedure provided by the manufacturer. Ultra-long DNA was extracted by the SDS method without purification step to sustain the length of DNA. The DNA degradation and contamination of the extracted DNA was monitored on 1% agarose gels. DNA purity was then detected using NanoDrop™ One UV-Vis spectrophotometer (Thermo Fisher Scientific, USA), of which OD260/280 ranging from 1.8 to 2.0 and OD 260/230 is between 2.0-2.2. At last, DNA concentration was further measured by Qubit® 4.0 Fluorometer (Invitrogen, USA).

### Nanopore sequencing and analysis

For each ultra-long Nanopore library, approximately 8-10 μg of gDNA was size-selected (>50 kb) with SageHLS HMW library system (Sage Science, USA), and processed using the Ligation sequencing 1D kit (SQK-LSK109, Oxford Nanopore Technologies, UK) according the manufacturer’s instructions. About 800ng DNA libraries were constructed and sequenced on the Promethion (Oxford Nanopore Technologies, UK) at the Genome Center of Grandomics (Wuhan, China). Base calling was performed with Guppy. QC metrics were calculated using pycoQC (https://github.com/a-slide/pycoQC). Reads were aligned to the chm13v2 reference genome^20^ with Winnowmap2^21^ due to superior representation and mapping of repetitive genomic sequence respectively. Reads that aligned to chromosome 4q and spanned the D4Z4 repeat array were retained for further analysis. We then used nucmer tool from the Mummer package to perform pairwise sequence alignment with the 3.3kb KpnI repeat element from GenBank sequence AF117653. Dotplots were generated with Dot (https://github.com/dnanexus/dot).

## Data Availability

All data and code are available on request. The raw sequence data has been deposited in the Genome Sequence Archive in National Genomics Data Center, China National Center for Bioinformation / Beijing Institute of Genomics, Chinese Academy of Sciences (GSA-Human: HRA003506) and are publicly accessible at https://ngdc.cncb.ac.cn/gsa-human.

## Acknowledgments

The authors thank participants for their contribution to this study.

## Author Contributions

KL, DQ, CES, PZ, and JGS designed the study. KL, FQ, YL, RH, AH, FL, RZ, SRD, YY, and WW collected data and performed experiments. KL, DQ, CES, PZ, and JGS analyzed data and wrote the manuscript. All authors approved this study prior to publication.

## Ethics Declaration

Elements of this study involving human subjects were approved by the Medical Ethics Committee of Beijing Tsinghua Changgung Hospital, Tsinghua University. All participants provided informed written consent before initiating research and data collection.

