## Supplemental Data for "Genetic Diagnosis of Facioscapulohumeral Muscular Dystrophy Type 1 Using Rare Variant Linkage Analysis and Long Read Genome Sequencing"

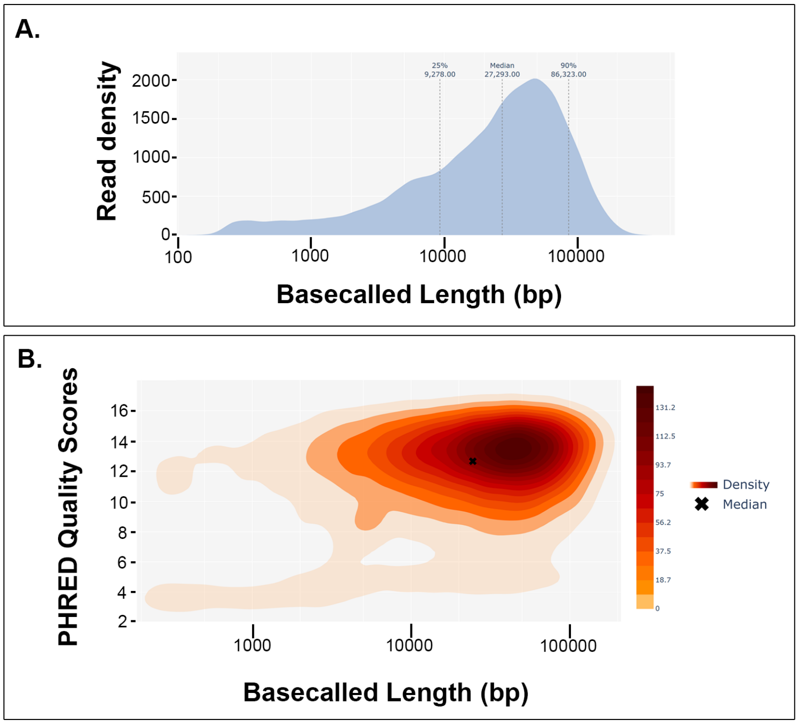


**Supplemental Figure 1. Sequencing metrics from Oxford Nanopore Sequencing.** (**A**) Read length distribution of all reads that pass quality metrics. Median, 95th percentile, and 25th percentile read lengths are noted. (**B**) Density plot of the relationship between sequencing read length and read quality score for all sequencing reads passing and failing quality metrics.

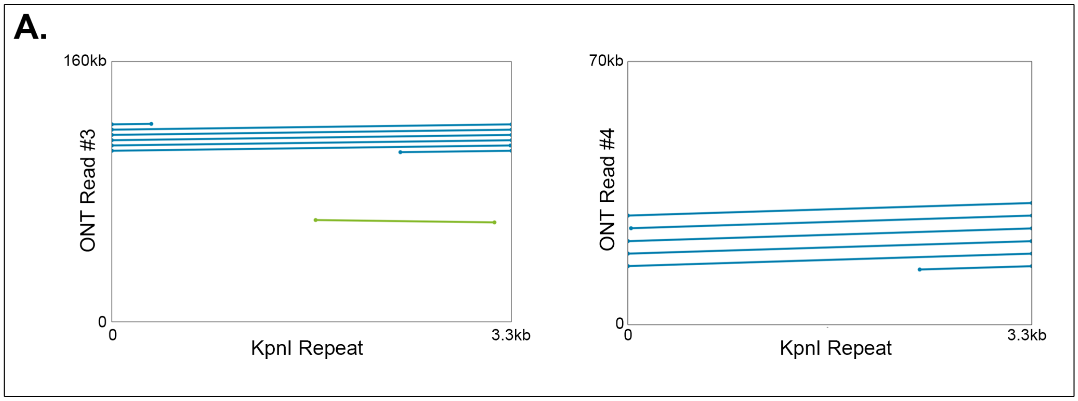


**Supplemental Figure 2. Additional nanopore reads supporting a pathogenic FSHD1 allele.** Sequence alignment of two additional nanopore sequencing reads (‘ONT Read 3’ and ‘ONT Read 4’) with the 3.3kb KpnI repeat sequence at D4Z4 locus demonstrate the presence of 5 KpnI repeats in an affected individual.

**Supplemental Table 1. Rare Variants in Linkage with the Disease Phenotype**

| Chromosome | RSID | Position | Reference Allele | Alternate Allele | Zygosity |
|---|---|---|---|---|---|
| chr4 | rs367635682 | 188855116 | T | C | Heterozygous |
| chr4 | rs138969910 | 188903131 | T | A | Heterozygous |
| chr4 | NA | 189894661 | C | T | Heterozygous |
| chr4 | rs910574891 | 189975788 | A | G | Heterozygous |
